# Effects of Hormone Therapy on survival, cancer, cardiovascular and dementia risks in 7 million menopausal women over age 65: a retrospective observational study

**DOI:** 10.1101/2022.05.25.22275595

**Authors:** Seo H. Baik, Fitsum Baye, Clement J. McDonald

## Abstract

**Background:** The long-term influence of menopausal hormone therapy remains unanswered due to the termination of randomized clinical trials and discordant findings from observational studies.

**Methods:** From 2007-2019 enrollment records of 100% Medicare beneficiaries, we identified 7 million female enrollees aged 65 or more. We identified type, route and strength of estrogen based on their prescription drug utilization records. Using vital status record and encounter records, we defined the first onset of thirteen patient outcomes; all-cause mortality; 5 cancers (breast, lung, endometrial, colorectal, ovarian cancers); 6 CV conditions (ischemic heart diseases, heart failure, venous thromboembolism, stroke, atrial fibrillation, acute myocardial infarction); and dementia. Then, we implemented an extended Cox regression analysis to examine the effects of type, route, and strength of estrogens on each of 13 study outcomes.

**Findings:** Estrogen monotherapy (ET) exhibited a significant, 20% (aHR=0.80; 95% CI 0.78-0.82), relative risk reduction of mortality. The reduction was greater with estradiol (aHR=0.78; 95% CI 0.75-0.80) than conjugated estrogen (aHR=0.86; 95% CI 0.85-0.88), and with vaginal (aHR=0.69; 95% CI 0.65-0.74) than oral (aHR=0.89; 95% CI 0.87-0.90) and transdermal (aHR=0.78; 95% CI 0.75-0.81) preparations. ET also exhibited significant risk *reductions* for all study cancers, breast (aHR=0.82; 95% CI 0.80-0.84), lung (aHR=0.87; 95% CI 0.84-0.90), endometrial (aHR=0.65; 95% CI 0.62-0.69), colorectal (aHR=0.86; 95% CI 0.82-0.90) and ovarian (aHR=0.83; 95% CI 0.79-0.88). ET slightly increased risks of ischemic heart diseases (aHR=1.03; 95% CI 1.01-1.04). However, such risk was not observed with low dose ET (aHR=0.98; 95% CI 0.97-0.99). Both combination therapy (aHR=1.11; 95% CI 1.08-1.14) and progestogen monotherapy (aHR=1.09; 95% CI 1.05-1.13) exhibited a significantly increased risk of breast cancer. Oral HT exhibited a moderately increased risk of dementia.

**Conclusions:** Among female Medicare beneficiaries aged ≥65, the effect of menopausal hormone therapy varies by type, route, and strength but overall estrogen seemed beneficial.

## Introduction

In 2002, the Women’s Health Initiatives (WHI) trial reported that menopausal estrogen+progestogen therapy (EPT) increased the occurrence of invasive breast cancer, stroke, and coronary heart disease in menopausal women (1) though it reduced fractures. The press dramatically presented the negative outcomes in a manner that some described as misleading (2,3). However, most of these results lost significance when corrected for multiple testing (1), and the second WHI study in 2004 examined the effect of estrogen monotherapy (ET) on these same outcomes and reported a near significant *reduction* in breast cancer (4), which became a significant 22% *reduction* in WHI’s long-term follow-up of that study (5). However, these positive results got little press attention and did little to reduce the fears about hormone therapy (HT) that had already implanted in the public’s mind. The WHI trials only studied one type (conjugated estrogen) and dosage (0.625 mg) and route (oral) of estrogen alone and in combination with a progestin. Many have wondered how different the results would have been with other types, routes, and dosage strengths of such medicines.

The U.S. Centers for Medicare and Medicaid Services (CMS) Virtual Research Data Center (VRDC) (6) carries 13 years of prescription claims and 20 years of encounter claims (7,8) as well as vital status for most US women over 65, and thus menopausal. So, VRDC provides HT exposure data as well as outcomes data about death, dementia, cardiovascular (CV), and cancer conditions like those studied by the WHI trials. Furthermore, at least 2-7% of elderly women still use HT after age 65 (9), enough that VRDC data might shed light on the consequences of HT use in older women and is large enough to provide data about the effect of dose size, routes and types of estrogens that the WHI trials could not. We implemented an extended Cox regression analyses (10–12) to assess the association of these factors with death, dementia, CV and cancer outcomes. Here we report the results of these analyses.

## Methods

### Study population

The CMS provided us access to all records of Medicare Part A (hospital insurance), B (medical insurance), and D (prescription drug insurance), claims data for 100% Part D enrollees. We constrained our study sample to be women who were first entitled in Medicare near age 65 (± 1 month) and during the full years of Part D benefits (i.e., 2007–2019) —a total of 13 years. We only included enrollees with at least 6 months of data –to assure enough follow-up time.

We report usage trends broken down by year, HT type and route, using the number of Part D female enrollees in each year as the denominator. We report descriptive statistics of patients’ demographics, socio-economic status, the prevalence of 49 chronic conditions, and the associated crude rates of outcomes.

### HT exposures

We classified HT into 6 related drug types, 4 routes and 3 dose strengths, as applicable. The drug types included estradiol (E2) alone, conjugated estrogen (CEE) alone, progestin (P) alone, E2+P combined, CEE+P combined and ethinyl estradiol (EE)+P combined. The routes include oral, transdermal, vaginal and injectable. We developed an average daily estrogen dose based on DailyMed (13) dosing instructions for each of 124 individual HT products (see full details in S1 Table) which accommodated intermittent regimens (e.g., 21 days on and 7 days off). We defined a “standard” dose for estrogen type and route, based on the literature and the distribution of daily estrogen doses as 0.625 mg, 1 mg, and 5 μg, for oral CEE, E2, and EE, respectively and 200 μg and 50 μg for non-oral, CEE and E2, respectively. For each drug type, we categorized this average daily estrogen doses into high, greater than 1.45 times the standard; low, less than 0.45 times the standard; and medium, between the lower end of the high, and the upper end of the low, bounds. We considered each combination of estrogen type, dose strength, and route as separate covariates (22 of them). We considered subjects to be exposed to a study drug if they ever had a prescription for that drug prior to an outcome event.

We only included HT medications with indications for menopausal symptoms and excluded the few indicated for birth control or for vaginal bleeding (injectable CEE). We did include injectable E2 which is indicated for menopausal symptoms (14,15). We did not include Megestrol in our study because of its special cancer uses.

### Outcomes

Our goals were to describe the usage of HT in women age ≥65 and determine the influence of such usage on survival, and on the occurrence of WHI-like outcomes: 5 cancers, 6 cardiovascular (CV) conditions, and dementia (1,4). The cancer outcomes included breast, lung, endometrial, colorectal, and ovarian, cancers. The CV outcomes included: ischemic heart diseases (IHD), heart failure (HF), venous thromboembolism (VTE), stroke, atrial fibrillation (AFB), and acute myocardial infarction (AMI). The occurrence and onset date for all but ovarian cancer and VTE were predefined by algorithm in Medicare’s Chronic Condition Data Warehouse (CCW) (16). We generated comparable occurrence data by examining ICD-9/10-CM codes for ovarian cancer (183.0 and C56) and VTE (415.1, 451, 453, I26, I80, I82), respectively.

### Statistical analysis

We explored the independent effect of each HT drug on the 13 outcomes listed above using separate extended Cox regression analyses. In each regression analysis, we included 22 combinations of HT drug type, routes, and dose ranges as covariates. As adjustments for patients’ characteristics, we also included race, degree of low-income-subsidy (LIS) (a surrogate for income (17)), rural residence indicator, calendar year of Medicare Part D enrollment (to adjust for secular trends), and the 47 CCW chronic conditions (16) with >1% prevalence, to adjust for overall medical burden. We treated all covariates except race as time-varying to avoid the risk of an immortal time bias and, protect against violations of proportional hazard assumption (10–12). When a cancer, or CV condition, or dementia was the outcome, we excluded all other cancer, CV conditions or dementia conditions, as covariates respectively.

Subjects became eligible for the study at the time of their Medicare entitlement, but prescription records were unavailable until their Part D enrollment. Many enrollees in our study also moved from Medicare Fee-For-Service (FFS) to a capitated plan (Medicare Advantage or MA) or disenrolled from Medicare. We had to censor them when that happened because we lost access to their encounter records and associated diagnose. We followed these subjects from their entry to Part D (while accounting for left truncation (18)) until they 1) experienced an outcome of interest, 2) switched to a capitated plan, 3) disenrolled from Medicare or 4) reached 12/31/2019 (the end of our data availability), whichever came first. In order to 1) mitigate selection bias toward HT use and to 2) correct for potential bias from informative censoring, we developed 2 *time-varying* propensity scores (PSs) using logistic regressions (19–21). The first was for the likelihood of taking HT and the second for the likelihood of switching from Medicare FFS to a capitated plan or disenrolling from Medicare altogether and thus dropping out of the study. Both PSs were conditional probabilities based on patient’s characteristics (demographics, socioeconomics, and presence of the 47 chronic conditions) and were iteratively estimated every 6 months among the patients who remained in follow-up considering all covariate values in a given 6-month cycle (21). We ran all Cox regression analyses with these time-varying PSs as additional adjustments (22,23).

### Secondary analysis

In this study, enrollees who moved from Medicare FFS to a capitated plan or disenrolled from Medicare had to be censored. Because such censoring could be *informative* and result in censoring bias (24), we included a PS of such drop-outs as an extra covariate in our primary analysis. In addition to that, we performed an intention-to-treat (ITT) analysis (25) to assess the possibility that censoring bias accounted for the difference in mortality risk associated with HT use from our primary analysis. In this secondary analysis, we included all female beneficiaries who were still on FFS at one year post Part D enrollment and followed them until death or the end of the study (12/31/2019) while ignoring dropouts after one year. The covariates in this secondary analysis include our study drugs, and all of the other covariates used in our primary analysis but were time-fixed as of their values at or before the one-year cut-off. In addition, we also include the PS of taking any HT as of the one-year cut-off (i.e., time-fixed). This ITT analysis would result in more accurate estimates of drug efficacy than ones yielded from a per-protocol analysis (26). We report the relative risk for each study drug from this analysis.

## Results

### Study population and Secular trends

From 100% Part D senior female enrollees, more than 7 million satisfied our selection criteria. The death cohort, our largest, included 7,036,466 individuals, and a modest proportion, 15%, of them used some type of HT at least once during our study period (Table 1). The disease-specific cohorts were slightly smaller in number but had similar proportions of HT users.

**Table 1.** Baseline Characteristics.

|  | **Any HT** | **Neither** |
| --- | --- | --- |
| **N** | 1,082,565 | 5,953,901 |
| **Age at Part D Entry, Median (IQR)** | 64.0(64.0-65.0) | 64.0(64.0-65.0) |
| **Age at The End of Follow-Up, Median (IQR)** | 71.0(68.0-74.0) | 69.0(67.0-72.0) |
| **White** | 965,436(89.2) | 4,791,602(80.5) |
| **Black** | 34,503(3.2) | 465,960(7.8) |
| **Hispanic** | 36,392(3.4) | 350,606(5.9) |
| **Asian** | 17,075(1.6) | 177,022(3.0) |
| **Other** | 29,159(2.7) | 168,711(2.8) |
| **Ever Dual** | 87,787(8.1) | 1,004,457(16.9) |
| **Non-Dual LIS** | 14,179(1.3) | 156,754(2.6) |
| **Non-Dual No LIS** | 980,599(90.6) | 4,792,690(80.5) |
| **Living In Rural Area** | 289,562(26.7) | 1,605,895(27.0) |
| **Hysterectomy** | 231,611(21.4) | 639,809(10.7) |
| **Cataract** | 718,924(66.4) | 2,954,232(49.6) |
| **Chronic Kidney Disease** | 221,782(20.5) | 1,284,284(21.6) |
| **COPD** | 174,424(16.1) | 932,907(15.7) |
| **Diabetes** | 240,201(22.2) | 1,664,431(28.0) |
| **Glaucoma** | 223,538(20.6) | 952,997(16.0) |
| **Hip/Pelvic Fracture** | 15,723(1.5) | 82,706(1.4) |
| **Depression** | 412,225(38.1) | 1,727,657(29.0) |
| **Alzheimer's Disease or Senile Dementia** | 63,605(5.9) | 309,437(5.2) |
| **Osteoporosis** | 268,653(24.8) | 1,135,562(19.1) |
| **Rheumatoid Arthritis/Osteoarthritis** | 657,040(60.7) | 2,638,202(44.3) |
| **Anemia** | 443,105(40.9) | 2,035,431(34.2) |
| **Asthma** | 167,973(15.5) | 667,100(11.2) |
| **Hyperlipidemia** | 811,559(75.0) | 4,016,394(67.5) |
| **Hypertension** | 721,052(66.6) | 3,926,742(66.0) |
| **Hypothyroidism** | 394,493(36.4) | 1,576,154(26.5) |
| **Alcohol Use Disorders** | 19,208(1.8) | 115,692(1.9) |
| **Anxiety Disorders** | 345,272(31.9) | 1,363,862(22.9) |
| **Bipolar Disorder** | 34,909(3.2) | 143,139(2.4) |
| **Major Depressive Affective Disorder** | 314,194(29.0) | 1,290,630(21.7) |
| **Drug Use Disorder** | 37,411(3.5) | 157,354(2.6) |
| **Personality Disorders** | 27,095(2.5) | 93,976(1.6) |
| **Schizophrenia and other Psychotic Disorders** | 18,016(1.7) | 107,525(1.8) |
| **Epilepsy** | 19,718(1.8) | 110,771(1.9) |
| **Cystic Fibrosis and other Metabolic Developmental Disorders** | 23,581(2.2) | 83,419(1.4) |
| **Fibromyalgia, Chronic Pain and Fatigue** | 439,070(40.6) | 1,569,200(26.4) |
| **Viral Hepatitis (General)** | 10,304(1.0) | 62,552(1.1) |
| **Liver Disease Cirrhosis and other Liver Conditions (Excluding Hepatitis)** | 105,312(9.7) | 487,132(8.2) |
| **Leukemias and Lymphomas** | 19,399(1.8) | 92,685(1.6) |
| **Migraine and other Chronic Headache** | 117,950(10.9) | 327,037(5.5) |
| **Mobility Impairments** | 20,922(1.9) | 140,658(2.4) |
| **Obesity** | 252,541(23.3) | 1,634,171(27.4) |
| **Overarching OUD Disorder** | 30,997(2.9) | 105,841(1.8) |
| **Peripheral Vascular Disease** | 112,913(10.4) | 601,917(10.1) |
| **Tobacco Use Disorders** | 78,084(7.2) | 633,390(10.6) |
| **Pressure Ulcers and Chronic Ulcers** | 35,812(3.3) | 221,553(3.7) |
| **Sensory - Deafness and Hearing Impairment** | 125,001(11.5) | 399,956(6.7) |

Note. Data are presented as No. (%) of patients unless otherwise noted.
Abbreviations: HT = Hormone Therapy; IQR = Interquartile Range; AMI = Acute Myocardial Infarction; COPD = Chronic Obstructive Pulmonary Disease.

Over the 13 years of follow-up (2007-2019), the proportion of senior women taking any HT containing estrogen dropped by half, from 11.4% to 5.8%. E2 tended to replace CEE; EPT plummeted from 1.4% to a minuscule 0.2% (Figure 1a); and vaginal route tended to replace oral route (Figure 1b). Overall, the number of ET users was 8 to 10 times greater than the number of EPT or P alone users, respectively. Among ET users, the predominant route was vagina. Twice as many women were on vaginal and 1/3rd as many on transdermal as on oral preparations. Among EPT users, the most common type of EPT was E2+P, followed by CEE+P and then EE+P (Table 2). All prescriptions of EE+P were a combination of EE and norethindrone and we did not include the 8% of the EE+P preparations indicated for birth control for consistency’s sake though that indication made no sense for our senior population.

**Figure 1.**
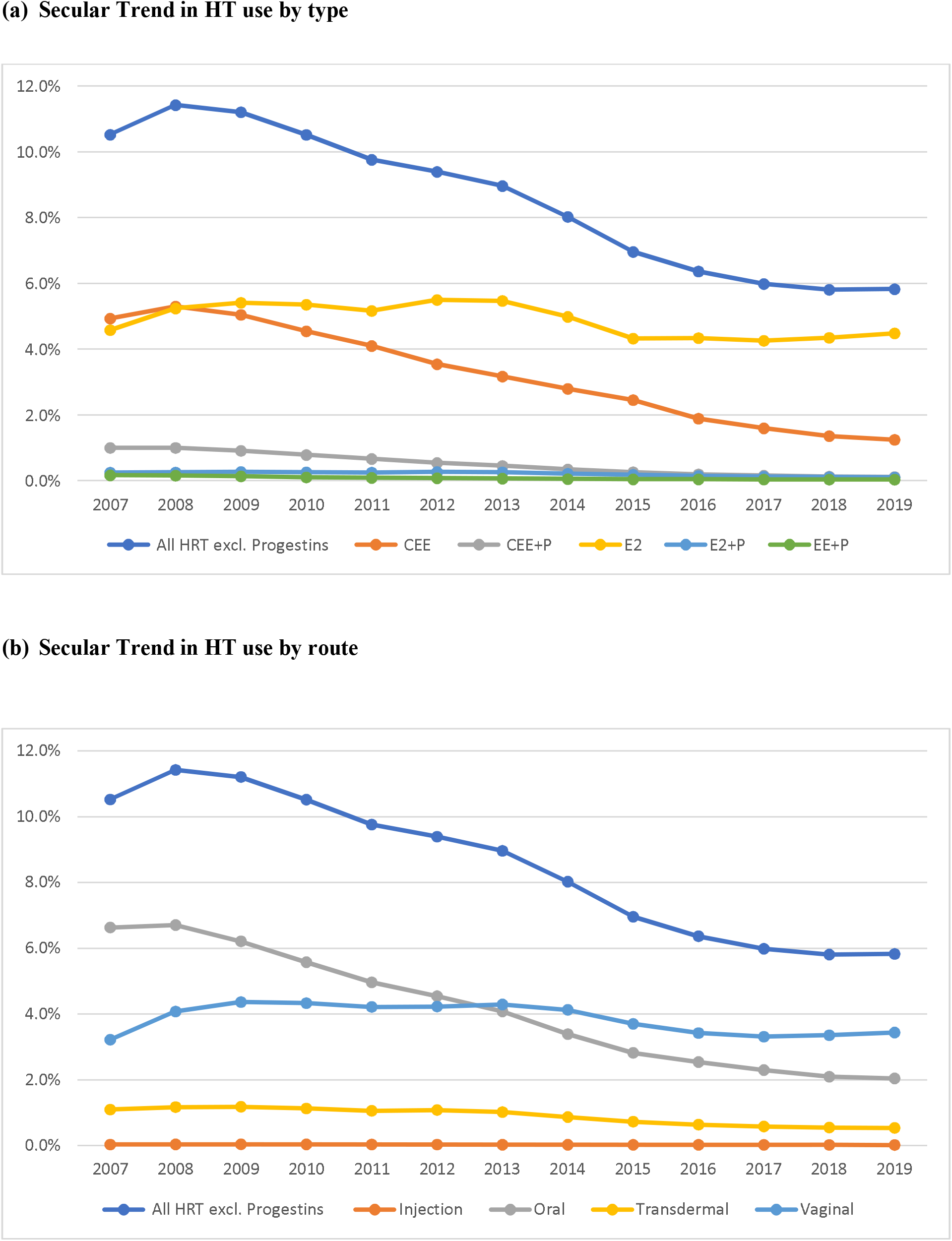
Trend in the use of Hormone Therapy (HT) by type and route.

**Table 2.**
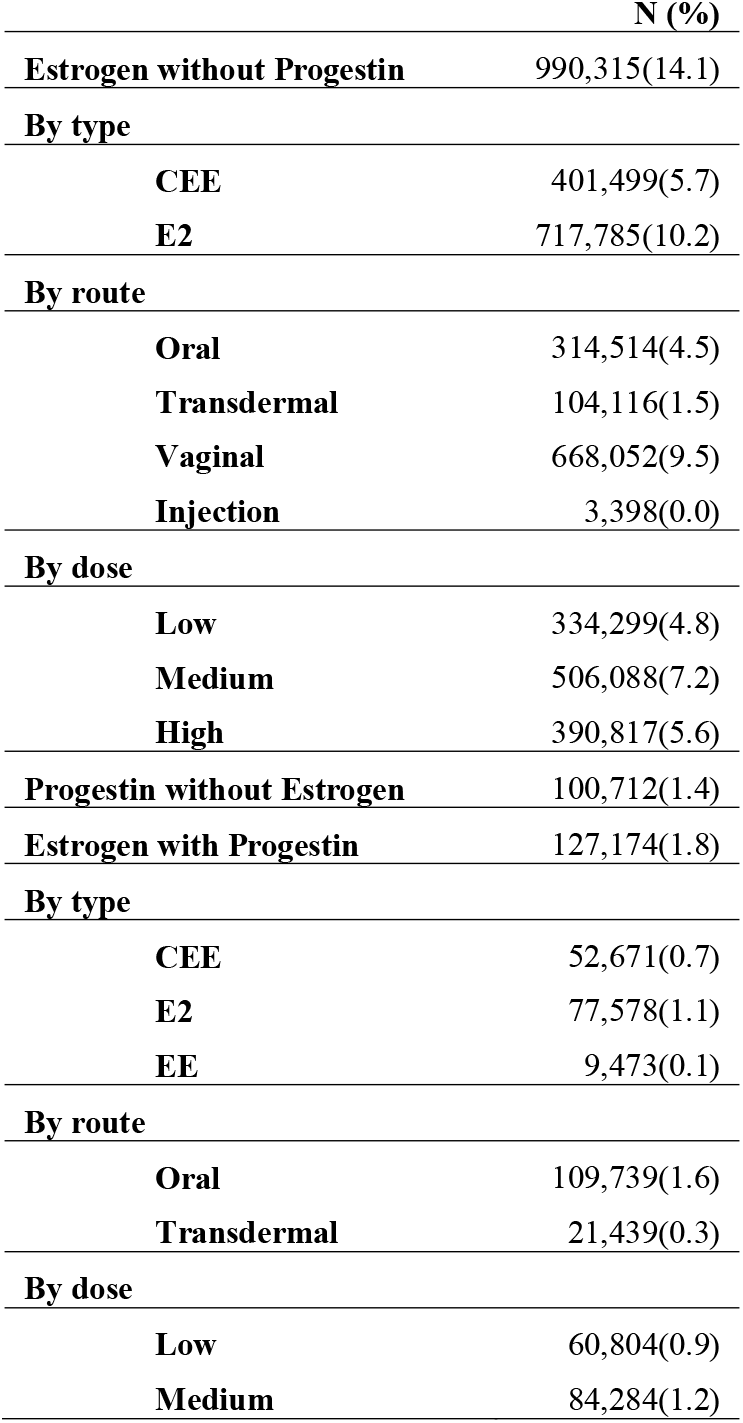
Number (Percentage) of patients by Route, Type, and Strength in Death Cohort (N = 7,036,466)

Starting with Part D enrollment, the median follow-up in death cohort was 4.1 years (total of 33,171,664 person-years); it was longer among any HT users (5.8 years) than no HT users (3.9 years) (Table 3). Overall, 384,376 patients died (5.5% or 12 per 1000 person-years). The death incidence was lower among HT users than no HT users (6 vs 13 per 1000 person-years). We censored 1.5 out of 7 million subjects as they left FFS for MA plans (17.7%) or dis-enrolled from Medicare (4.5%). The rate of censoring for this reason was higher among no HT users, indicating potential bias from informative censoring. The number of subjects and follow-up duration varied somewhat across disease-specific cohorts because of different end points and censoring rates by outcomes.

**Table 3.**
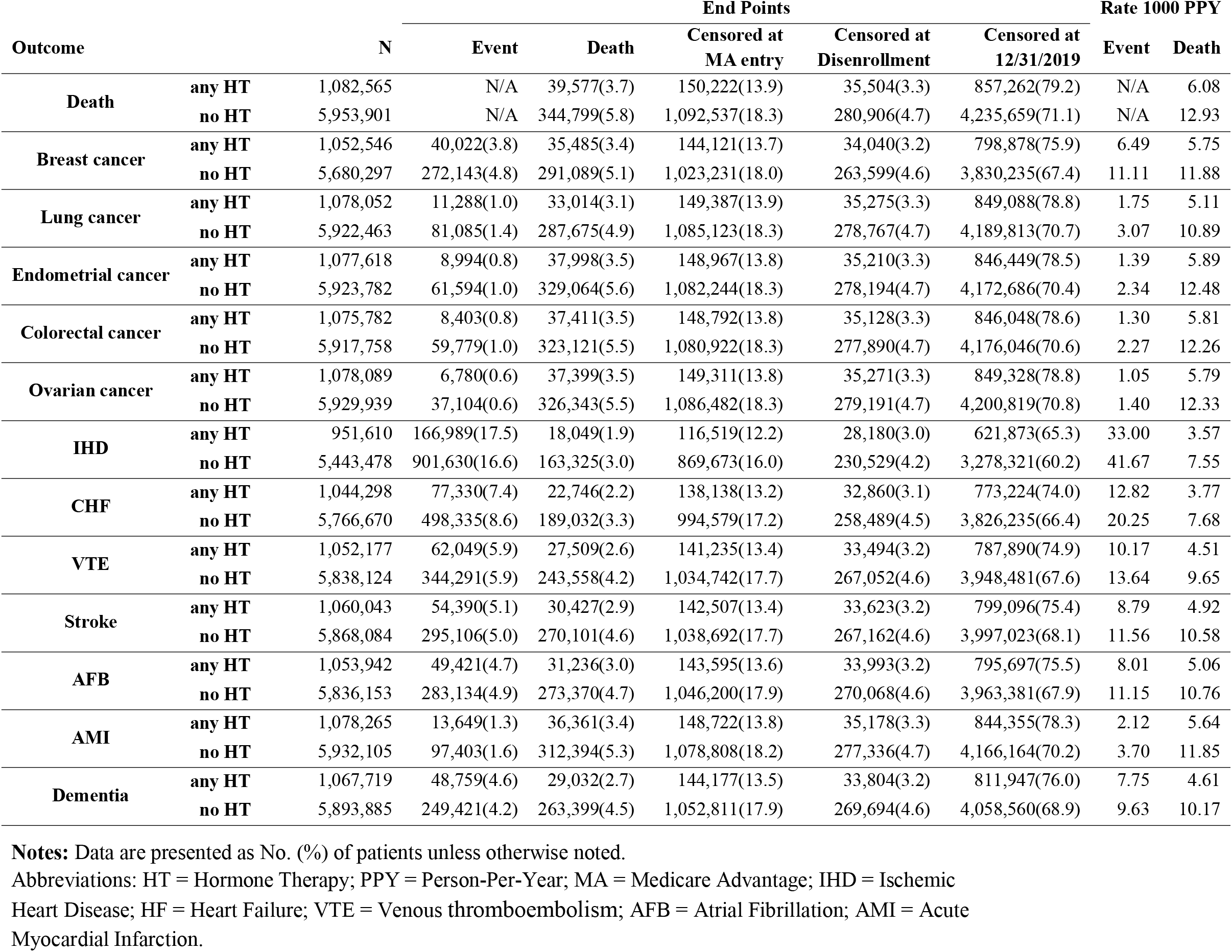
Event/Censoring Points and Rates of Event/Death by Each Study Cohort.

Medicaid eligibility for special supplements was our proxy for income level. Accordingly, there were three income groups; dual (15%) with incomes below 135% of the Federal Poverty Line (FPL); non-dual LIS (2.4%) between 135% and 150% FPL; and non-dual no LIS (82.0%) above 150% FLP (27). The proportions of non-Hispanic White and rural resident were 81.8% and 26.9% respectively. Among the chronic conditions, hyperlipidemia (68.6%), hypertension (66.1%), and cataract (52.2%), were the most common (Table 1). Because we had no claims data about hysterectomies performed before age 65, hysterectomy data was only available for 12.4% of our cohort and most of it came from the ICD diagnosis codes for “acquired absence of uterus/cervix” (S2 Table).

### Primary Analyses

In Tables 4 and 5, we present the marginal relative risk of 13 study outcomes associated with the use of each HT after controlling for all time-varying and time-fixed covariates including the use of any other HT beyond the index HT, namely adjusted hazard ratio (aHR). We report aHR as the percent of risk above (increased) or below (decreased) one by an amount of 100 x (aHR-1)%. We present marginal aHR by overall use of ET, EPT and Progestin to highlight average differences by type, route, and dose level. We also present aHR for each of 22 combinations of type, route, and dose level in Tables 6 and 7, where oral medium dose CEE and CEE+P indicate comparable drugs studied in the WHI trials.

**Table 4.**
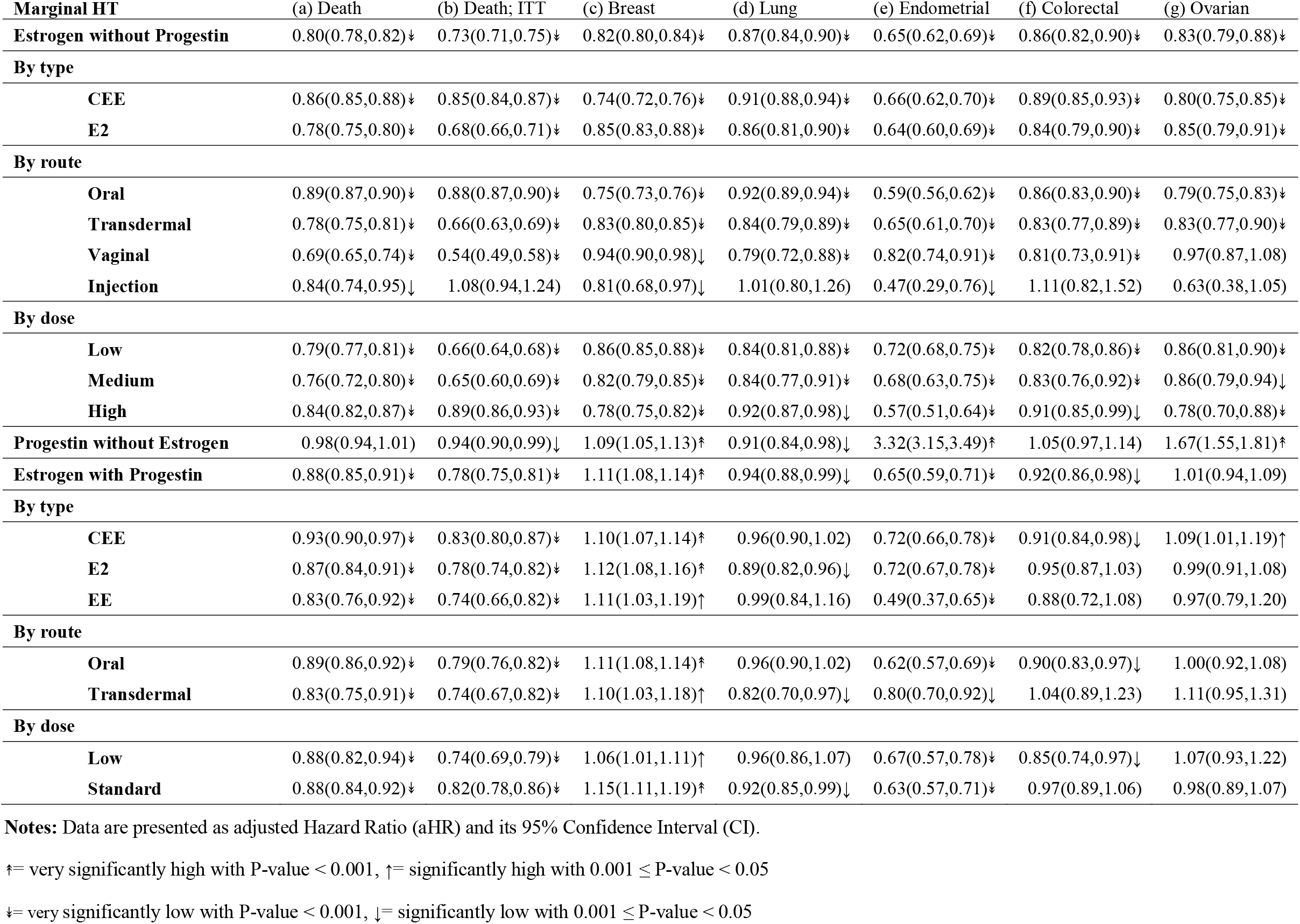
Marginal effects of HT regimens on all-cause mortality and cancer outcomes.

**Table 5.** Marginal effects of HT regimens on CV outcomes and dementia.

| Marginal HT | (a) IHD | (b) CHF | (c) VTE | (d) Stroke | (e) AFM | (f) AMI | (g) Dementia |
| --- | --- | --- | --- | --- | --- | --- | --- |
| **Estrogen without Progestin** | 1.03(1.02,1.04)† | 0.95(0.94,0.96)‡ | 0.96(0.95,0.98)‡ | 1.01(1.00,1.03) | 0.95(0.93,0.97)‡ | 0.89(0.86,0.93)‡ | 0.98(0.96,0.99)↓ |
| **By type** |  |  |  |  |  |  |  |
| CEE | 1.04(1.03,1.05)† | 0.99(0.98,1.00) | 0.97(0.96,0.99)↓ | 1.06(1.04,1.08)† | 0.99(0.98,1.01) | 0.93(0.90,0.96)‡ | 1.01(0.99,1.03) |
| E2 | 1.03(1.02,1.05)† | 0.93(0.92,0.95)‡ | 0.96(0.94,0.98)‡ | 1.00(0.98,1.02) | 0.93(0.91,0.95)‡ | 0.88(0.84,0.92)‡ | 0.96(0.94,0.99)↓ |
| **By route** |  |  |  |  |  |  |  |
| Oral | 1.03(1.02,1.04)† | 1.01(0.99,1.02) | 0.97(0.96,0.99)‡ | 1.06(1.05,1.08)† | 0.99(0.98,1.01) | 0.93(0.91,0.96)‡ | 1.01(1.00,1.03)↑ |
| Transdermal | 1.00(0.98,1.01) | 0.89(0.87,0.91)‡ | 0.93(0.91,0.95)‡ | 0.94(0.92,0.97)‡ | 0.92(0.89,0.94)‡ | 0.87(0.82,0.92)‡ | 0.91(0.89,0.94)‡ |
| Vaginal | 1.03(1.00,1.05)↑ | 0.87(0.84,0.90)‡ | 0.96(0.92,0.99)↓ | 0.98(0.94,1.02) | 0.93(0.89,0.97)↓ | 0.83(0.75,0.91)‡ | 0.94(0.90,0.99)↓ |
| Injection | 1.19(1.10,1.28)† | 1.17(1.06,1.29)↑ | 1.02(0.91,1.15) | 1.12(0.99,1.27) | 0.86(0.74,0.99)↓ | 1.05(0.85,1.31) | 1.08(0.95,1.22) |
| **By dose** |  |  |  |  |  |  |  |
| Low | 0.98(0.97,0.99)‡ | 0.86(0.85,0.88)‡ | 0.92(0.91,0.94)‡ | 0.94(0.92,0.95)‡ | 0.93(0.91,0.95)‡ | 0.83(0.80,0.86)‡ | 0.91(0.89,0.93)‡ |
| Medium | 1.02(1.00,1.04)↑ | 0.93(0.90,0.96)‡ | 0.96(0.93,0.99)↓ | 1.00(0.97,1.04) | 0.95(0.92,0.99)↓ | 0.88(0.82,0.95)↓ | 0.98(0.94,1.01) |
| High | 1.09(1.07,1.11)† | 1.05(1.03,1.08)† | 1.00(0.97,1.03) | 1.09(1.06,1.13)† | 0.96(0.93,0.99)↓ | 0.96(0.91,1.01) | 1.03(1.00,1.06)↑ |
| **Progestin without Estrogen** | 1.03(1.01,1.05)↑ | 1.04(1.01,1.07)↑ | 1.03(1.00,1.06) | 0.99(0.95,1.02) | 1.04(1.01,1.08)↑ | 0.86(0.81,0.93)‡ | 0.99(0.95,1.03) |
| **Estrogen with Progestin** | 0.96(0.94,0.97)‡ | 0.89(0.87,0.91)‡ | 0.96(0.93,0.98)↓ | 0.93(0.90,0.95)‡ | 0.92(0.89,0.95)‡ | 0.93(0.87,0.98)↓ | 1.03(1.00,1.06)↑ |
| **By type** |  |  |  |  |  |  |  |
| CEE | 0.95(0.93,0.96)‡ | 0.91(0.89,0.94)‡ | 0.94(0.91,0.97)‡ | 0.93(0.90,0.96)‡ | 0.92(0.89,0.95)‡ | 0.95(0.90,1.01) | 1.02(0.98,1.05) |
| E2 | 0.98(0.96,1.00)↓ | 0.89(0.86,0.92)‡ | 0.96(0.93,0.99)↓ | 0.93(0.90,0.96)‡ | 0.92(0.89,0.96)‡ | 0.95(0.88,1.02) | 1.04(1.00,1.07) |
| EE | 0.93(0.89,0.97)↓ | 0.87(0.81,0.94)‡ | 0.97(0.91,1.05) | 0.92(0.85,1.00) | 0.91(0.84,0.99)↓ | 0.87(0.73,1.03) | 1.04(0.95,1.12) |
| **By route** |  |  |  |  |  |  |  |
| Oral | 0.94(0.93,0.96)‡ | 0.89(0.87,0.91)‡ | 0.96(0.93,0.98)↓ | 0.93(0.90,0.96)‡ | 0.91(0.88,0.94)‡ | 0.93(0.87,0.99)↓ | 1.03(1.00,1.07)↑ |
| Transdermal | 1.03(1.00,1.07) | 0.91(0.85,0.97)↓ | 0.96(0.90,1.02) | 0.93(0.87,1.00) | 0.98(0.91,1.05) | 0.90(0.77,1.05) | 1.01(0.94,1.09) |
| **By dose** |  |  |  |  |  |  |  |
| Low | 0.94(0.91,0.97)‡ | 0.87(0.83,0.92)‡ | 0.95(0.90,1.00)↓ | 0.90(0.85,0.96)‡ | 0.89(0.84,0.95)‡ | 0.88(0.78,0.98)↓ | 0.97(0.91,1.02) |
| Standard | 0.97(0.95,0.99)↓ | 0.91(0.88,0.93)‡ | 0.96(0.93,0.99)↓ | 0.95(0.91,0.98)↓ | 0.94(0.90,0.97)↓ | 0.97(0.90,1.04) | 1.08(1.04,1.12)† |

**Notes:** Data are presented as adjusted Hazard Ratio (aHR) and its 95% Confidence Interval (CI).
Abbreviations: IHD = ischemic heart diseases; HF = heart failure; VTE = venous thromboembolism; AFB = atrial fibrillation; AMI = acute myocardial infarction.
↑= very significantly high with P-value < 0.001, ↑= significantly high with $0.001 \leq \text{P-value} < 0.05$
↓= very significantly low with P-value < 0.001, ↓= significantly low with $0.001 \leq \text{P-value} < 0.05$

**Table 6.**
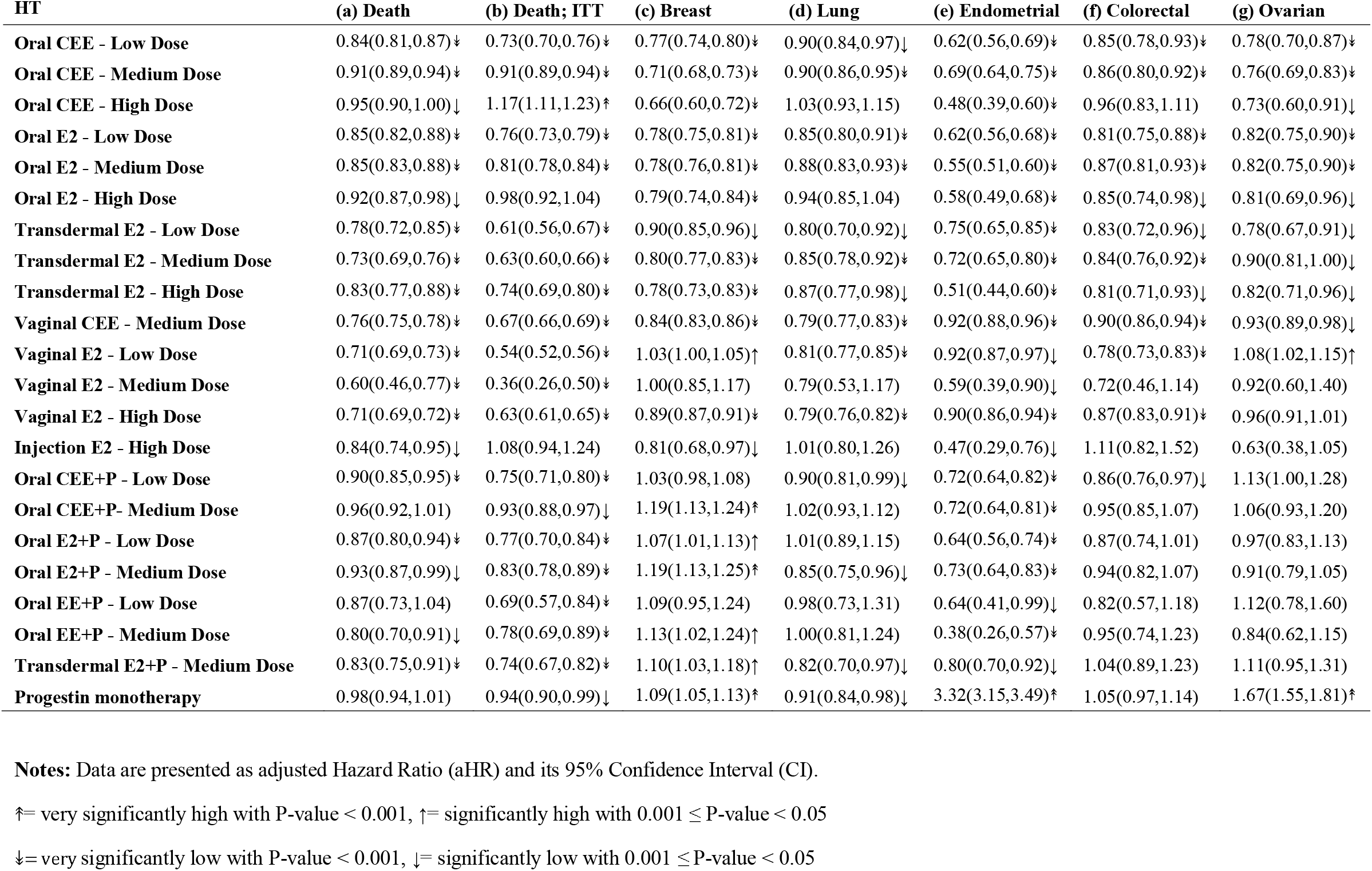
Hazard ratios of 22 HT regimens on all-cause mortality and cancer outcomes.

**Table 7.**
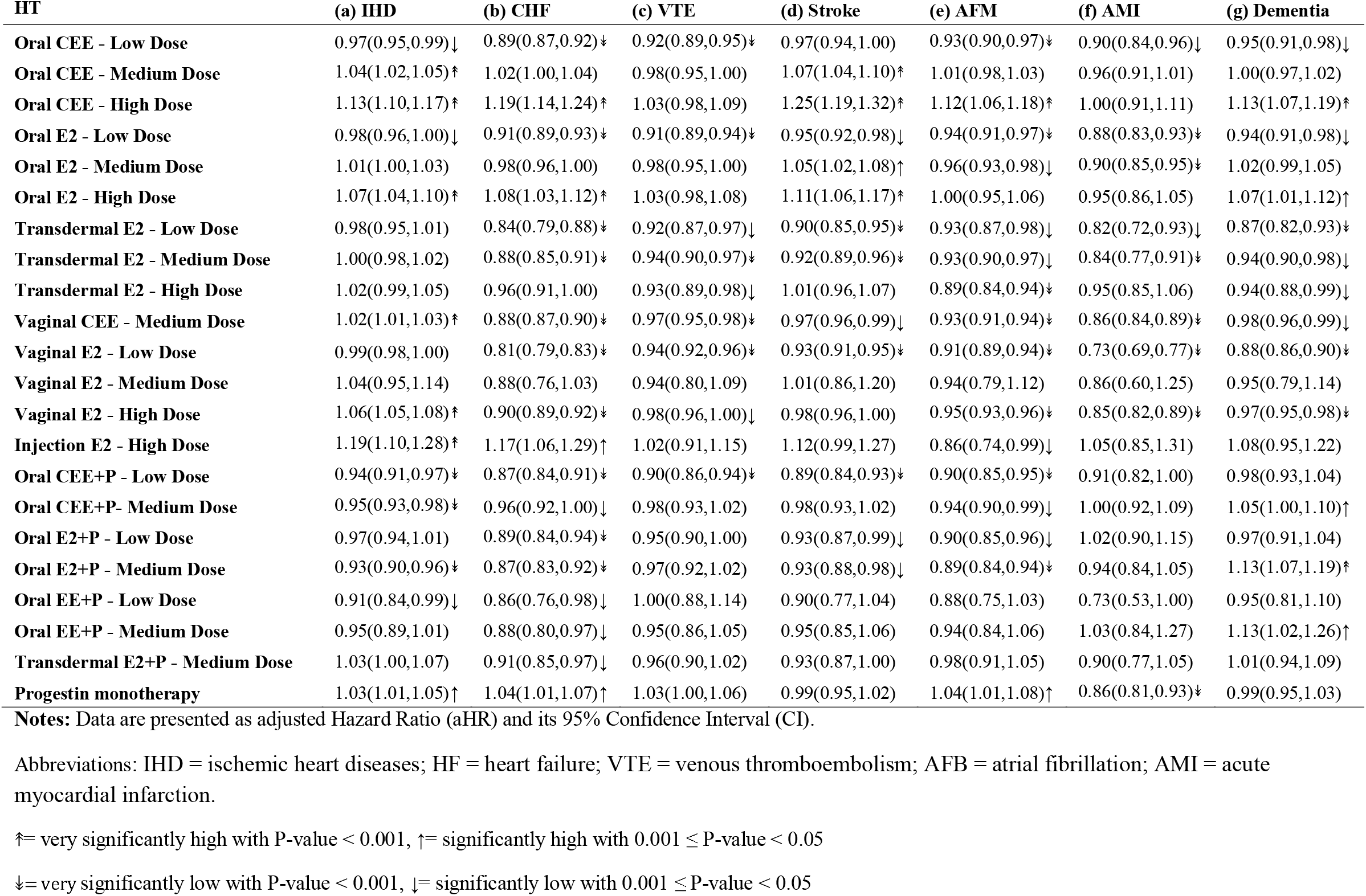
Hazard ratios of 22 HT regimens on CV outcomes and dementia.

### The risk of all-cause mortality associated with HT use

On average, ET use was associated with a significant, 20% reduction in mortality risk relative to no ET use (Table 4-a) which translated to 77,401 fewer expected deaths in our large population. All combinations of ET type, route, and dose were also associated with reduced mortality risk. The marginal risk of E2 on mortality was significantly less than that of CEE. Vaginal, transdermal, and oral preparation, in order by size of mortality risk, each had significantly less mortality risk than the one after. The mortality risk of low and medium doses were significantly less than high dose but were not different from each other. Overall, EPT was associated with significant 12% reduction in marginal risk of mortality. Progestin monotherapy had no significant association. Interestingly, oral CEE medium dose, comparable to the drug in the WHI trial of ET, exhibited less risk reduction in mortality (9%) than overall ET (Table 6-a).

The 6,405,685 patients (90% of those in the primary analysis) still in the study 12 months after Part D enrollment were the subjects for our ITT analysis. No censoring occurred after the 12-month cut-off in the ITT analysis; so, could not have shaped these results. Yet, marginal association between each type, route and dose level of HT and death from this analysis paralleled and exceeded the mortality reduction observed in the primary analysis (Table 3-b). So, the biases due to informative censoring are unlikely to account for the mortality reduction observed in the primary analysis.

### The risk of breast, lung, endometrial, colorectal, and ovarian, cancer associated with HT use

During our study period, breast cancer incidence was four times that of any other study cancer (Table 3). ET use was associated with significant *reduction* in marginal risk of breast cancer, 18% overall, as well as within each combination of ET type, route, and dose size (Table 4-c). Oral ET exhibited significantly greater reductions than transdermal and transdermal ET significantly greater than vaginal. Furthermore, CEE was associated with significantly greater risk reduction for breast cancer than E2 – a pattern opposite to what we saw with mortality as the outcome. The intervention in WHI’s 13-year long-term follow-up postintervention study, oral CEE 0.625 mg, was associated with a 21% % risk reduction for breast cancer (5). Seventy percent of our oral CEE medium dose prescriptions were 0.625 mg, same strength as the WHI dose, 11% were greater (0.9 mg) and 19% less (0.45 mg) than 0.625 mg. In our study, oral CEE medium dose use was associated with a 29% risk reduction and was the second best among all oral ET (Table 6-c). To the negative, both EPT and progestin monotherapy significantly *increased* the risk of breast cancer (Table 3-c).

ET was associated with 13% and 14% decrease in the risks of lung and colorectal cancers, respectively. EPT also exhibited reductions in lung and colorectal cancer risks but by smaller amounts than ET. Progestin monotherapy was also associated with reduced lung cancer risk but had no association with colorectal cancer risk (Table 3-d & f).

With ET use, endometrial, and ovarian cancer risks declined moderately (Table 3-e & g) but this finding is likely an artifact of the selective use of ET in hysterectomized women who lack the organs where such cancers could arise. More than half of hysterectomized women also had bilateral oophorectomy (28). So, these two findings might not be meaningful. On the other hand, 35% risk reduction of endometrial cancer associated with EPT use could be meaningful because it is usually prescribed for women with an intact uterus.

### The risk of IHD, CHF, VTE, Stroke, AFM, AMI, and Dementia, risk associated with HT

IHD occurred in more than a million subjects, twice the size of the next most frequent CV condition, CHF. So, ET’s 3% increased risk of IHD is of overweight importance. Most types, routes and dose levels of ET were also associated with increased risk of IHD, up to 19% with the injectables, but less so (3-4%) in other types, route, and doses. Specifically, low dose oral CEE and E2 were associated with significant risk reduction for IHD by 3% and 2%, respectively (Table 7-a). Progestin monotherapy had the same 3% increased risk for IHD as ET. Interestingly, EPT formulations, overall, were associated with a significant 4% risk reduction of IHD (Table 4-a).

To the positive, ET use, overall, was associated with 5% decrease *in* the risk of CHF (Table 4-b). However, among the ET subgroups of high dose and injectable ET were associated with increased risk of CHF. As was true for IHD, all EPT formulations were associated with *reduced* CHF risk.

Overall ET use had small or no, associations with, stroke or dementia, risk (Table 4-d & g). However, high and medium doses of oral E2 and CEE had important increased risk of stroke, reaching 25% with high dose oral CEE (Table 7-d & g). Both estrogen types (CEE and E2) were also associated with increased dementia risk but only with high dose oral preparations.

Transdermal and vaginal preparations of ET, that avoid first pass travel through the liver were associated with small *reduce*d or null risk of both conditions, consistent with the results of other studies (29) and with the procoagulant and proinflammation effects of estrogen’s liver passage (30,31). EPT use, on average, was also associated with small but significant, *increased* risk of dementia and, decreased risk of all 6 CV conditions.

## Discussion

This study is a population-based retrospective observational study that explored the associations between estrogen types, routes, and dose size and all-cause mortality, dementia, 5 common cancers, and 6 cardiovascular, conditions using an extended Cox regression analysis. Important limitations included that data availability only began at age 65; hysterectomy information was unavailable for most of our subjects.; we depended on claims for encounter diagnoses and could not validate them through chart review; and, as is true for all observational studies, differential influences of unmeasured confounders, such as adherence to healthy behavior among HT users, could have been present.

One strength was the use of filled prescription records, rather than patient recall, to ascertain HT use. Another strength was its size —a study population of more than 7 million menopausal women, nearly an order of magnitude larger than any previous HT study (32). Its large size let us estimate differences in associations between each of 22 HT drug type, route and dose combinations on the one hand and the many study outcomes on the other.

### Association with all-cause mortality

ET use, overall, was associated with a 20% reduction in mortality risk considering all ET preparations together. The sole estrogen preparation studied in the WHI trials was 0.625 mg of oral CEE, and it exhibited a close to significant, 6% (HR=0.94; 95% CI 0.88-1.01) mortality reduction in the 18-year long-term follow-up of the WHI trails (33). This result gives some plausibility to the 9% mortality reduction we observed in association with the use of our medium dose, oral CEE which was almost identical to the drug randomized in WHI’s estrogen only trial. In our study, vaginal E2 in low and medium doses were associated with greatest reduction of mortality risk (29-40%) (Table 6-a). Our overall mortality results are consistent with the mortality results from an meta-analysis of 31 observational and RCT studies that reported reduced mortality among HT users (34) and with the re-analyses of the Prostate, Lung, Colorectal, and Ovarian (PLCO) Cancer Screening RTC, which reported a 23% decrease in all-cause mortality among current users of any HT (35).

### Associations with cancer

In our study, overall ET use and oral CEE medium dose use were associated with an 18% and 29% reduction of breast cancer risk, respectively. The long-term follow-up of the WHI study reported a significant, and similarly sized, 21% reduction in breast cancer incidence (36) giving credence to this result.

In our study, ET use was also associated with significantly reduced colorectal (14%) and lung (13%) cancer risk as was EPT, but to a lesser degree. A few observational studies support our observations of reduced lung cancer rates in association with HT use (37,38), though there is controversy. Two observational studies (39,40) and a re-analysis of the PLCO trial data (35) support our protective associations between HT use and colorectal cancer. The greater incidence of colorectal (+79%) (41) and lung cancer (+26%) (42) among patients with lower level of estrogen due to oophorectomy as part of their hysterectomy compared to hysterectomized patients without ovary removal also support our results by implying that estrogen protects against these two cancers. On the downside, both EPT and progestin alone were associated with significant *increases* (11% and 9% respectively) in breast cancer risk.

### Associations with CV disease and dementia

Our data revealed a 3% increase in risk for IHD with ET use compared to no use. However, this increased risk is concentrated in the use of high (9%) and medium (2%) dose levels. Low dose ET, used by one third of ET users, exhibited a salubrious, 2%, d*ecrease*d risk for IHD. The associations between ET use and other CV conditions was also to the good—4-11% risk reductions. E2 and vaginal/transdermal compared to CEE and oral preparation exhibited numerically lower risks for all CV conditions. It further suggests that a similar decrease in dementia risk would accrue when low dose, topical (vaginal, transdermal) and E2, were prescribed and in stroke risk when low dose topicals and low dose oral E2 were prescribed. Topical ET preparations in our study, that would avoid the procoagulant and proinflammatory effects ascribed to liver passage (30,31), were associated with *reduce*d risk of both conditions, consistent with the results of other studies (29).

### Overall considerations

The risk reduction associated with various HT we saw generally favor less use of EPT, more use of the topical (vaginal or transdermal) rather than the oral route, use E2 rather than CEE, and low or medium dose rather than high dose for menopausal care. These advantages have been emphasized by others (43) and our trend data suggest prescribers have adopted them to some degree. However, our results on breast cancer are less consistent with patterns we saw with mortality as the outcome. ET use overall exhibited a decreased risk of breast cancer, but the reduction was significantly greater among CEE, oral route, and high dose, uses, than their counter parts. Further our EPT was used only by small subset of HT users (12%) and in that subset its use was associated with a reduction in the risk of endometrial cancer.

We did not include fractures as an outcome in our study. However, among the outcomes we included, we saw significant association with the use of specific HT medications that paralleled effects seen in each of the *significant* WHI end points including, the decreased incidence of colorectal cancer with both ET and EPT use (35), the decreased incidence of breast cancer with ET use, the increased incidence of breast cancer with EPT respectively and the decreased incidence of endometrial cancer with EPT use (36). These parallels give credence to our observational results.

Our follow-up began when women entered Medicare at about age 65, but it is likely that many of them started taking HT closer to the time of their menopausal symptoms and continued it into their Medicare years. If so, our positive results align with the timing hypotheses (44) that asserts HT use early in menopauses is better than later, but extend it by reporting positive effects with usage continued into Medicare years. Such use is not a rarity per our data.

The FDA’s black box warning about HT (45) extrapolates from the deleterious effect of EPT on breast cancer (1) and dementia (46) to all forms of ET, because no studies to the contrary existed then. Now, such studies do exist. The long-term WHI studies reported a significant decrease in breast cancer incidence (5) and death due to Alzheimer’s disease (33), among patient in the ET study arm. Furthermore, many studies suggest that E2, topical routes, and low doses may produce better outcomes than CEE, oral and medium dose (0.625 mg) studied in the WHI’s ET study (47,48).

Menopausal HT of all types has been maligned by the results of earliest WHI study (1), long public memory of the frightening media reports about it (2) and lack of attention to recent more positive studies. Our study though purely observational raises the *possibility* of important improvements in survival and reduction in the risk of three cancers. We hope our results stimulate, interest in hormone therapy and further observational studies using other large national database and biologic research, to support them.

## Data Availability

All data produced in the present work are contained in the manuscript

## Acknowledgement

The authors thank Terry Therneau (PhD, Professor of Biostatistics, Mayo Clinic) for his guiding our use of time varying Cox regression to this data set and especially for the idea of using Intention to treat approach to identify biases due to censoring.

## Contributorship Statement

SHB and CJM conceived and designed the study. SHB and FB performed the statistical analysis. SHB and CJM drafted the manuscript and all authors contributed substantially to the study.

## Data Availability Statement

The Medicare claims data belong to CMS, and we do not have permission to publish the raw data. However, the data are available to researchers through the CMS Virtual Research Data Center.

We have included almost all study results in our tables and supporting information.

## Supporting information

**Description of S1 Table**

**S1 Table. All menopausal hormone therapy estrogen and progestin drugs available in Medicare prescription drug claims data during our 13-year study**.

**S2 Table. List of CPT AND ICD Diagnosis and Procedure codes used to define the absence of a uterus or hysterectomy surgery**.

