## Supplementary Tables for "Effects of Hormone Therapy on survival, cancer, cardiovascular and dementia risks in 7 million menopausal women over age 65: a retrospective observational study"

**Description of S1 Table**

### We listed every pure estrogen, combination E+P, and pure P, product prescribed during our study in S1 Table.

This table includes a row for each unique combination of HT drug (generic, brand) name, route, and strength that we observed over 13 years of Medicare claims. We describe each column’s contents in order by its column number in the following.

(1) The HT product’s generic name
(2) brand name
(3) medication route type, i.e., oral, transdermal, vaginal or injectable.
(4) form, i.e., tablet, cream, gel, spray, patch, ring etc.
(5) strength string which will include multiple strength, when a produce includes multiple distinct ingredients). Cells in this column also include a hyperlink to the website from which we obtained the product’s label information. One can click on this link to see the full label.
(6) Available average daily estrogen doses in µg. When the label described more than one dosing, more than one average daily dose will appear in this column, separated by semicolons.
(7) Categorization of the estrogen strength of that adjusted dose as low, medium, or high.

**S1 Table. All menopausal hormone therapy estrogen and progestin drugs available in Medicare prescription drug claims data during our 13-year study.**

| **Generic Name** | **Brand Name** | **Route** | **Form** | **Strength** | **Average Daily Dose (µg)** | **Strength Class** |
| --- | --- | --- | --- | --- | --- | --- |
| CEE | Premarin | Oral | Tablet | [0.3 MG](https://dailymed.nlm.nih.gov/dailymed/drugInfo.cfm?setid=258e1602-a3cf-4ccc-ca80-73dbbfb812ff) | 250; 300 | Low |
| CEE | Premarin | Oral | Tablet | [0.45MG](https://dailymed.nlm.nih.gov/dailymed/drugInfo.cfm?setid=258e1602-a3cf-4ccc-ca80-73dbbfb812ff) | 375; 450 | Medium |
| CEE | Premarin | Oral | Tablet | [0.625 MG](https://dailymed.nlm.nih.gov/dailymed/drugInfo.cfm?setid=258e1602-a3cf-4ccc-ca80-73dbbfb812ff) | 521; 625 | Medium |
| CEE | Premarin | Oral | Tablet | [0.9 MG](https://dailymed.nlm.nih.gov/dailymed/drugInfo.cfm?setid=258e1602-a3cf-4ccc-ca80-73dbbfb812ff) | 750; 900 | Medium |
| CEE | Premarin | Oral | Tablet | [1.25 MG](https://dailymed.nlm.nih.gov/dailymed/drugInfo.cfm?setid=258e1602-a3cf-4ccc-ca80-73dbbfb812ff) | 1042; 1250 | High |
| CEE | Premarin | Vaginal | Cream | [0.625 MG/G](https://dailymed.nlm.nih.gov/dailymed/drugInfo.cfm?setid=96609623-528e-4aba-cabe-7254aed816d5#S2.1) | 89; 234 | Medium |
| CEE-Medroxyprogesterone | Premphase | Oral | Tablet | [0.625 (14)](https://dailymed.nlm.nih.gov/dailymed/drugInfo.cfm?setid=fd0c0836-5d23-2183-da81-9dc7f4287052) | 625 | Medium |
| CEE-Medroxyprogesterone | Prempro | Oral | Tablet | [0.3-1.5 MG](https://dailymed.nlm.nih.gov/dailymed/drugInfo.cfm?setid=fd0c0836-5d23-2183-da81-9dc7f4287052) | 300 | Low |
| CEE-Medroxyprogesterone | Prempro | Oral | Tablet | [0.45-1.5 MG](https://dailymed.nlm.nih.gov/dailymed/drugInfo.cfm?setid=fd0c0836-5d23-2183-da81-9dc7f4287052) | 450 | Medium |
| CEE-Medroxyprogesterone | Prempro | Oral | Tablet | [0.625-2.5](https://dailymed.nlm.nih.gov/dailymed/drugInfo.cfm?setid=fd0c0836-5d23-2183-da81-9dc7f4287052) | 625 | Medium |
| CEE-Medroxyprogesterone | Prempro | Oral | Tablet | [0.625-5 MG](https://dailymed.nlm.nih.gov/dailymed/drugInfo.cfm?setid=fd0c0836-5d23-2183-da81-9dc7f4287052) | 625 | Medium |
| E2 | Estrace | Oral | Tablet | [0.5 MG](https://dailymed.nlm.nih.gov/dailymed/drugInfo.cfm?setid=a3803ba3-4eee-4e2e-ac8c-821a4e6720cc) | 500 | Low |
| E2 | Estrace | Oral | Tablet | [1 MG](https://dailymed.nlm.nih.gov/dailymed/drugInfo.cfm?setid=a3803ba3-4eee-4e2e-ac8c-821a4e6720cc) | 1000 | Medium |
| E2 | Estrace | Oral | Tablet | [2 MG](https://dailymed.nlm.nih.gov/dailymed/drugInfo.cfm?setid=a3803ba3-4eee-4e2e-ac8c-821a4e6720cc) | 2000 | High |
| E2 | Estradiol | Oral | Tablet | [0.5 MG](https://dailymed.nlm.nih.gov/dailymed/drugInfo.cfm?setid=a3803ba3-4eee-4e2e-ac8c-821a4e6720cc) | 500 | Low |
| E2 | Estradiol | Oral | Tablet | [1 MG](https://dailymed.nlm.nih.gov/dailymed/drugInfo.cfm?setid=a3803ba3-4eee-4e2e-ac8c-821a4e6720cc) | 1000 | Medium |
| E2 | Estradiol | Oral | Tablet | [2 MG](https://dailymed.nlm.nih.gov/dailymed/drugInfo.cfm?setid=a3803ba3-4eee-4e2e-ac8c-821a4e6720cc) | 2000 | High |
| E2 | Gynodiol | Oral | Tablet | [1.5 MG](https://www.drugs.com/imprints/0158-logo-10704.html) | 1500 | High |
| E2 | Gynodiol | Oral | Tablet | [1MG](https://www.drugs.com/imprints/1259-logo-8368.html) | 1000 | Medium |
| E2 | Alora | Transdermal | Patch | [.025MG/24H](https://dailymed.nlm.nih.gov/dailymed/drugInfo.cfm?setid=ac676026-14be-486a-9d64-6702bd9e51ea) | 25 | Low |
| E2 | Alora | Transdermal | Patch | [.075MG/24H](https://dailymed.nlm.nih.gov/dailymed/drugInfo.cfm?setid=ac676026-14be-486a-9d64-6702bd9e51ea) | 75 | Medium |
| E2 | Alora | Transdermal | Patch | [0.05MG/24H](https://dailymed.nlm.nih.gov/dailymed/drugInfo.cfm?setid=ac676026-14be-486a-9d64-6702bd9e51ea) | 50 | Medium |
| E2 | Alora | Transdermal | Patch | [0.1MG/24HR](https://dailymed.nlm.nih.gov/dailymed/drugInfo.cfm?setid=ac676026-14be-486a-9d64-6702bd9e51ea) | 100 | High |
| E2 | Climara | Transdermal | Patch | [.025MG/24H](https://dailymed.nlm.nih.gov/dailymed/drugInfo.cfm?setid=1e9702c4-f2d7-4ea8-b6e8-7dca31671864) | 25 | Low |
| E2 | Climara | Transdermal | Patch | [.0375MG/24](https://dailymed.nlm.nih.gov/dailymed/drugInfo.cfm?setid=1e9702c4-f2d7-4ea8-b6e8-7dca31671864) | 37 | Medium |
| E2 | Climara | Transdermal | Patch | [.075MG/24H](https://dailymed.nlm.nih.gov/dailymed/drugInfo.cfm?setid=1e9702c4-f2d7-4ea8-b6e8-7dca31671864) | 75 | Medium |
| E2 | Climara | Transdermal | Patch | [0.05MG/24H](https://dailymed.nlm.nih.gov/dailymed/drugInfo.cfm?setid=1e9702c4-f2d7-4ea8-b6e8-7dca31671864) | 50 | Medium |
| E2 | Climara | Transdermal | Patch | [0.06MG/24H](https://dailymed.nlm.nih.gov/dailymed/drugInfo.cfm?setid=1e9702c4-f2d7-4ea8-b6e8-7dca31671864) | 60 | Medium |
| E2 | Climara | Transdermal | Patch | [0.1MG/24HR](https://dailymed.nlm.nih.gov/dailymed/drugInfo.cfm?setid=1e9702c4-f2d7-4ea8-b6e8-7dca31671864) | 100 | High |
| E2 | Divigel | Transdermal | Gel | [0.25(0.1%)](https://dailymed.nlm.nih.gov/dailymed/drugInfo.cfm?setid=59e610dc-883e-4f07-9ec6-a5841b0bf9fb) | 250 | High |
| E2 | Divigel | Transdermal | Gel | [0.5MG(0.1)](https://dailymed.nlm.nih.gov/dailymed/drugInfo.cfm?setid=59e610dc-883e-4f07-9ec6-a5841b0bf9fb) | 500 | High |
| E2 | Divigel | Transdermal | Gel | [0.75/0.75G](https://dailymed.nlm.nih.gov/dailymed/drugInfo.cfm?setid=59e610dc-883e-4f07-9ec6-a5841b0bf9fb) | 750 | High |
| E2 | Divigel | Transdermal | Gel | [1MG(0.1%)](https://dailymed.nlm.nih.gov/dailymed/drugInfo.cfm?setid=59e610dc-883e-4f07-9ec6-a5841b0bf9fb) | 1000 | High |
| E2 | Dotti | Transdermal | Patch | [.025MG/24H](https://dailymed.nlm.nih.gov/dailymed/drugInfo.cfm?setid=53016d30-6b5f-49da-ac16-85ef415a5805) | 25 | Low |
| E2 | Dotti | Transdermal | Patch | [.0375MG/24](https://dailymed.nlm.nih.gov/dailymed/drugInfo.cfm?setid=53016d30-6b5f-49da-ac16-85ef415a5805) | 37 | Medium |
| E2 | Dotti | Transdermal | Patch | [.075MG/24H](https://dailymed.nlm.nih.gov/dailymed/drugInfo.cfm?setid=53016d30-6b5f-49da-ac16-85ef415a5805) | 75 | Medium |
| E2 | Dotti | Transdermal | Patch | [0.05MG/24H](https://dailymed.nlm.nih.gov/dailymed/drugInfo.cfm?setid=53016d30-6b5f-49da-ac16-85ef415a5805) | 50 | Medium |
| E2 | Dotti | Transdermal | Patch | [0.1MG/24HR](https://dailymed.nlm.nih.gov/dailymed/drugInfo.cfm?setid=53016d30-6b5f-49da-ac16-85ef415a5805) | 100 | High |
| E2 | Elestrin | Transdermal | Gel | [0.87G](https://dailymed.nlm.nih.gov/dailymed/drugInfo.cfm?setid=eff2dea1-f117-11e3-ac10-0800200c9a66) | 520 | High |
| E2 | Esclim | Transdermal | Patch | [0.05MG/24H](https://www.webmd.com/drugs/2/drug-17767/esclim-transdermal/details) | 50 | Medium |
| E2 | Estraderm | Transdermal | Patch | [0.05MG/24H](https://www.accessdata.fda.gov/drugsatfda_docs/label/2012/019081s042lbl.pdf) | 37; 50 | Medium |
| E2 | Estraderm | Transdermal | Patch | [0.1MG/24HR](https://www.accessdata.fda.gov/drugsatfda_docs/label/2012/019081s042lbl.pdf) | 75; 100 | High |
| E2 | Estradiol | Transdermal | Patch | [.025MG/24H](https://dailymed.nlm.nih.gov/dailymed/drugInfo.cfm?setid=e7e6da3b-8485-1382-61c9-e9b369018b98) | 25 | Low |
| E2 | Estradiol | Transdermal | Patch | [.0375MG/24](https://dailymed.nlm.nih.gov/dailymed/drugInfo.cfm?setid=e7e6da3b-8485-1382-61c9-e9b369018b98) | 37 | Medium |
| E2 | Estradiol | Transdermal | Patch | [.075MG/24H](https://dailymed.nlm.nih.gov/dailymed/drugInfo.cfm?setid=e7e6da3b-8485-1382-61c9-e9b369018b98) | 75 | Medium |
| E2 | Estradiol | Transdermal | Patch | [0.05MG/24H](https://dailymed.nlm.nih.gov/dailymed/drugInfo.cfm?setid=e7e6da3b-8485-1382-61c9-e9b369018b98) | 50 | Medium |
| E2 | Estradiol | Transdermal | Patch | [0.06MG/24H](https://dailymed.nlm.nih.gov/dailymed/drugInfo.cfm?setid=e7e6da3b-8485-1382-61c9-e9b369018b98) | 60 | Medium |
| E2 | Estradiol | Transdermal | Patch | [0.1MG/24HR](https://dailymed.nlm.nih.gov/dailymed/drugInfo.cfm?setid=e7e6da3b-8485-1382-61c9-e9b369018b98) | 100 | High |
| E2 | Estrasorb | Transdermal | Emulsion | [2.5/G-1.74](https://www.rxlist.com/estrasorb-drug.htm#indications) | 50 | Medium |
| E2 | Estrogel | Transdermal | Gel | [1.25G](https://dailymed.nlm.nih.gov/dailymed/drugInfo.cfm?setid=87bb0e2f-9fa6-438b-9d5f-d0a90af770ed) | 750 | High |
| E2 | Evamist | Transdermal | Spray | [1.53/SPRAY](https://dailymed.nlm.nih.gov/dailymed/drugInfo.cfm?setid=9a0aa631-133d-406b-9d32-8a1a99af4e50) | 1530 | High |
| E2 | Menostar | Transdermal | Patch | [14MCG/24HR](https://dailymed.nlm.nih.gov/dailymed/drugInfo.cfm?setid=a78708cd-04d4-4221-a4e4-53680bbe2912) | 14 | Low |
| E2 | Minivelle | Transdermal | Patch | [.025MG/24H](https://dailymed.nlm.nih.gov/dailymed/drugInfo.cfm?setid=6c5c47ab-28ee-11e1-bfc2-0800200c9a66) | 25 | Low |
| E2 | Minivelle | Transdermal | Patch | [.0375MG/24](https://dailymed.nlm.nih.gov/dailymed/drugInfo.cfm?setid=6c5c47ab-28ee-11e1-bfc2-0800200c9a66) | 37 | Medium |
| E2 | Minivelle | Transdermal | Patch | [.075MG/24H](https://dailymed.nlm.nih.gov/dailymed/drugInfo.cfm?setid=6c5c47ab-28ee-11e1-bfc2-0800200c9a66) | 75 | Medium |
| E2 | Minivelle | Transdermal | Patch | [0.05MG/24H](https://dailymed.nlm.nih.gov/dailymed/drugInfo.cfm?setid=6c5c47ab-28ee-11e1-bfc2-0800200c9a66) | 50 | Medium |
| E2 | Minivelle | Transdermal | Patch | [0.1MG/24HR](https://dailymed.nlm.nih.gov/dailymed/drugInfo.cfm?setid=6c5c47ab-28ee-11e1-bfc2-0800200c9a66) | 100 | High |
| E2 | Vivelle | Transdermal | Patch | [.025MG/24H](https://dailymed.nlm.nih.gov/dailymed/drugInfo.cfm?setid=e6f2e7ed-41a9-4a53-ac80-0cb1fa2edcbf) | 18; 25 | Low |
| E2 | Vivelle | Transdermal | Patch | [.0375MG/24](https://dailymed.nlm.nih.gov/dailymed/drugInfo.cfm?setid=e6f2e7ed-41a9-4a53-ac80-0cb1fa2edcbf) | 28; 37 | Medium |
| E2 | Vivelle | Transdermal | Patch | [0.05MG/24H](https://dailymed.nlm.nih.gov/dailymed/drugInfo.cfm?setid=e6f2e7ed-41a9-4a53-ac80-0cb1fa2edcbf) | 37; 50 | Medium |
| E2 | Vivelle | Transdermal | Patch | [0.1MG/24HR](https://dailymed.nlm.nih.gov/dailymed/drugInfo.cfm?setid=e6f2e7ed-41a9-4a53-ac80-0cb1fa2edcbf) | 75; 100 | High |
| E2 | Vivelle-Dot | Transdermal | Patch | [.025MG/24H](https://dailymed.nlm.nih.gov/dailymed/drugInfo.cfm?setid=e6f2e7ed-41a9-4a53-ac80-0cb1fa2edcbf) | 18; 25 | Low |
| E2 | Vivelle-Dot | Transdermal | Patch | [.0375MG/24](https://dailymed.nlm.nih.gov/dailymed/drugInfo.cfm?setid=e6f2e7ed-41a9-4a53-ac80-0cb1fa2edcbf) | 28; 37 | Medium |
| E2 | Vivelle-Dot | Transdermal | Patch | [.075MG/24H](https://dailymed.nlm.nih.gov/dailymed/drugInfo.cfm?setid=e6f2e7ed-41a9-4a53-ac80-0cb1fa2edcbf) | 56; 75 | Medium |
| E2 | Vivelle-Dot | Transdermal | Patch | [0.05MG/24H](https://dailymed.nlm.nih.gov/dailymed/drugInfo.cfm?setid=e6f2e7ed-41a9-4a53-ac80-0cb1fa2edcbf) | 37; 50 | Medium |
| E2 | Vivelle-Dot | Transdermal | Patch | [0.1MG/24HR](https://dailymed.nlm.nih.gov/dailymed/drugInfo.cfm?setid=e6f2e7ed-41a9-4a53-ac80-0cb1fa2edcbf) | 75; 100 | High |
| E2 | Estrace | Vaginal | Cream | [0.01%](https://dailymed.nlm.nih.gov/dailymed/drugInfo.cfm?setid=dfc41fcd-c5ba-4b25-b647-cf04d3988b3e) | 150; 225; 300 | High |
| E2 | Estradiol | Vaginal | Cream | [0.01%](https://dailymed.nlm.nih.gov/dailymed/drugInfo.cfm?setid=c94738ff-dece-4fb6-bb44-4f8832a45f38) | 150; 225; 300 | High |
| E2 | Estring | Vaginal | Ring | [7.5MCG/24H](https://dailymed.nlm.nih.gov/dailymed/drugInfo.cfm?setid=aa530dfd-3a48-46b9-9678-a7bc48316e41) | 7 | Low |
| E2 | Imvexxy | Vaginal | Insert | [4 MCG](https://dailymed.nlm.nih.gov/dailymed/drugInfo.cfm?setid=104be9f2-a8f6-430e-9e01-2ee7cc1861f1) | 3 | Low |
| E2 | Estradiol | Vaginal | Tablet | [10 MCG](https://dailymed.nlm.nih.gov/dailymed/drugInfo.cfm?setid=86620f6c-fd02-40df-98d5-3048dcd92c37) | 7 | Low |
| E2 | Imvexxy | Vaginal | Insert | [10 MCG](https://dailymed.nlm.nih.gov/dailymed/drugInfo.cfm?setid=104be9f2-a8f6-430e-9e01-2ee7cc1861f1) | 7 | Low |
| E2 | Vagifem | Vaginal | Tablet | [10 MCG](https://dailymed.nlm.nih.gov/dailymed/drugInfo.cfm?setid=e5ad3cf6-dd96-4e64-af21-c1eee38d0b88) | 7 | Low |
| E2 | Vagifem | Vaginal | Tablet | [25 MCG](https://dailymed.nlm.nih.gov/dailymed/drugInfo.cfm?setid=e5ad3cf6-dd96-4e64-af21-c1eee38d0b88) | 19 | Low |
| E2 | Yuvafem | Vaginal | Tablet | [10 MCG](https://dailymed.nlm.nih.gov/dailymed/drugInfo.cfm?setid=d20abc9a-8f47-e274-c9b1-b71c9489761f) | 7 | Low |
| E2 Acetate | Femtrace | Oral | Tablet | [0.45MG](https://www.accessdata.fda.gov/drugsatfda_docs/label/2014/021633s005lbl.pdf) | 450 | Low |
| E2 Acetate | Femtrace | Oral | Tablet | [0.9MG](https://www.accessdata.fda.gov/drugsatfda_docs/label/2014/021633s005lbl.pdf) | 900 | Medium |
| E2 Acetate | Femtrace | Oral | Tablet | [1.8MG](https://www.accessdata.fda.gov/drugsatfda_docs/label/2014/021633s005lbl.pdf) | 1800 | High |
| E2 Acetate | Femring | Vaginal | Ring | [0.05MG/24H](https://dailymed.nlm.nih.gov/dailymed/drugInfo.cfm?setid=7aaa97f9-a9d1-c815-e053-2991aa0afb9e) | 50 | Medium |
| E2 Acetate | Femring | Vaginal | Ring | [0.1MG/24HR](https://dailymed.nlm.nih.gov/dailymed/drugInfo.cfm?setid=7aaa97f9-a9d1-c815-e053-2991aa0afb9e) | 100 | High |
| E2 Cypionate | Depo-Estradiol | Injection | Vial | [5 MG/ML](https://dailymed.nlm.nih.gov/dailymed/drugInfo.cfm?setid=9a4229fd-fecd-4ac1-9c4f-6d442533457f) | 178; 35 | High |
| E2 Valerate | Delestrogen | Injection | Vial | [10 MG/ML](https://dailymed.nlm.nih.gov/dailymed/drugInfo.cfm?setid=e8f94df9-0692-4462-a7f0-fe019a0a3f07) | 357 | High |
| E2 Valerate | Delestrogen | Injection | Vial | [20 MG/ML](https://dailymed.nlm.nih.gov/dailymed/drugInfo.cfm?setid=e8f94df9-0692-4462-a7f0-fe019a0a3f07) | 714 | High |
| E2 Valerate | Delestrogen | Injection | Vial | [40 MG/ML](https://dailymed.nlm.nih.gov/dailymed/drugInfo.cfm?setid=e8f94df9-0692-4462-a7f0-fe019a0a3f07) | 2857; 2142 | High |
| E2 Valerate | Estradiol Valerate | Injection | Vial | [10 MG/ML](https://dailymed.nlm.nih.gov/dailymed/drugInfo.cfm?setid=e8f94df9-0692-4462-a7f0-fe019a0a3f07) | 357 | High |
| E2 Valerate | Estradiol Valerate | Injection | Vial | [20 MG/ML](https://dailymed.nlm.nih.gov/dailymed/drugInfo.cfm?setid=e8f94df9-0692-4462-a7f0-fe019a0a3f07) | 714 | High |
| E2 Valerate | Estradiol Valerate | Injection | Vial | [40 MG/ML](https://dailymed.nlm.nih.gov/dailymed/drugInfo.cfm?setid=e8f94df9-0692-4462-a7f0-fe019a0a3f07) | 2857; 2142 | High |
| E2-Drospirenone | Angeliq | Oral | Tablet | [0.25-0.5 MG](https://dailymed.nlm.nih.gov/dailymed/drugInfo.cfm?setid=761834c2-6b61-4583-84c2-f1ca4a97c4f2) | 500 | Low |
| E2-Drospirenone | Angeliq | Oral | Tablet | [0.5 MG-1 MG](https://dailymed.nlm.nih.gov/dailymed/drugInfo.cfm?setid=761834c2-6b61-4583-84c2-f1ca4a97c4f2) | 1000 | Medium |
| E2-Levonorgestrel | Climara Pro | Transdermal | Patch | [45-15/24H](https://dailymed.nlm.nih.gov/dailymed/drugInfo.cfm?setid=184d3092-7fc6-4375-816b-1ab06bb99cfd) | 45 | Medium |
| E2-Norethindrone | Activella | Oral | Tablet | [0.5-0.1 MG](https://dailymed.nlm.nih.gov/dailymed/drugInfo.cfm?setid=32a9f9a3-2940-251a-e054-00144ff88e88) | 500 | Low |
| E2-Norethindrone | Activella | Oral | Tablet | [1 MG-0.5 MG](https://dailymed.nlm.nih.gov/dailymed/drugInfo.cfm?setid=32a9f9a3-2940-251a-e054-00144ff88e88) | 1000 | Medium |
| E2-Norethindrone | Amabelz | Oral | Tablet | [0.5-0.1 MG](https://dailymed.nlm.nih.gov/dailymed/drugInfo.cfm?setid=3b8a7426-6f1c-470a-ac36-b8465b8679cd) | 500 | Low |
| E2-Norethindrone | Amabelz | Oral | Tablet | [1 MG-0.5MG](https://dailymed.nlm.nih.gov/dailymed/drugInfo.cfm?setid=3b8a7426-6f1c-470a-ac36-b8465b8679cd) | 1000 | Medium |
| E2-Norethindrone | Estradiol-Norethindrone Acetat | Oral | Tablet | [0.5-0.1 MG](https://dailymed.nlm.nih.gov/dailymed/drugInfo.cfm?setid=6cafe148-bfb8-4c09-a056-b0637c0c3065) | 500 | Low |
| E2-Norethindrone | Estradiol-Norethindrone Acetat | Oral | Tablet | [1 MG-0.5MG](https://dailymed.nlm.nih.gov/dailymed/drugInfo.cfm?setid=6cafe148-bfb8-4c09-a056-b0637c0c3065) | 1000 | Medium |
| E2-Norethindrone | Lopreeza | Oral | Tablet | [0.5-0.1 MG](https://dailymed.nlm.nih.gov/dailymed/drugInfo.cfm?setid=bf0e39cb-95e7-4831-92d2-0b3f071aeaae) | 500 | Low |
| E2-Norethindrone | Lopreeza | Oral | Tablet | [1 MG-0.5MG](https://dailymed.nlm.nih.gov/dailymed/drugInfo.cfm?setid=bf0e39cb-95e7-4831-92d2-0b3f071aeaae) | 1000 | Medium |
| E2-Norethindrone | Mimvey | Oral | Tablet | [1 MG-0.5MG](https://dailymed.nlm.nih.gov/dailymed/drugInfo.cfm?setid=b1c5c06b-9ce2-4bf7-9693-e37608055a14) | 1000 | Medium |
| E2-Norethindrone | Mimvey Lo | Oral | Tablet | [0.5-0.1 MG](https://dailymed.nlm.nih.gov/dailymed/drugInfo.cfm?setid=b1c5c06b-9ce2-4bf7-9693-e37608055a14) | 500 | Low |
| E2-Norethindrone | Combipatch | Transdermal | Patch | [.05-.14/24](https://dailymed.nlm.nih.gov/dailymed/drugInfo.cfm?setid=83198ef1-11c4-11e4-9191-0800200c9a66) | 50 | Medium |
| E2-Norethindrone | Combipatch | Transdermal | Patch | [.05-.25/24](https://dailymed.nlm.nih.gov/dailymed/drugInfo.cfm?setid=83198ef1-11c4-11e4-9191-0800200c9a66) | 50 | Medium |
| E2-Norgestimate | Ortho-Prefest | Oral | Tablet | [1-1-0.09MG](https://dailymed.nlm.nih.gov/dailymed/drugInfo.cfm?setid=70d00600-b8d3-4820-8db6-40d4536a6f9e) | 1000 | Medium |
| E2-Norgestimate | Prefest | Oral | Tablet | [1-1-0.09MG](https://dailymed.nlm.nih.gov/dailymed/drugInfo.cfm?setid=70d00600-b8d3-4820-8db6-40d4536a6f9e) | 1000 | Medium |
| E2-Progesterone | Bijuva | Oral | Capsule | [1MG-100MG](https://dailymed.nlm.nih.gov/dailymed/drugInfo.cfm?setid=bc52489a-5149-411d-9665-2c75405ad15d) | 1000 | Medium |
| EE-Norethindrone | Femhrt | Oral | Tablet | [0.5MG-2.5](https://www.accessdata.fda.gov/drugsatfda_docs/label/2005/21065s012lbl.pdf) | 2.5 | Low |
| EE-Norethindrone | Femhrt | Oral | Tablet | [1MG-5MCG](https://dailymed.nlm.nih.gov/dailymed/drugInfo.cfm?setid=0fece0aa-2b9c-40a3-975e-1ef2c2ef72f2) | 5 | Medium |
| EE-Norethindrone | Fyavolv | Oral | Tablet | [0.5MG-2.5](https://dailymed.nlm.nih.gov/dailymed/drugInfo.cfm?setid=772c86d7-10a4-48c5-8c19-f8aac68f5ab4) | 2 | Low |
| EE-Norethindrone | Fyavolv | Oral | Tablet | [1MG-5MCG](https://dailymed.nlm.nih.gov/dailymed/drugInfo.cfm?setid=772c86d7-10a4-48c5-8c19-f8aac68f5ab4) | 5 | Medium |
| EE-Norethindrone | Jevantique | Oral | Tablet | [1MG-5MCG](https://www.drugs.com/pro/jevantique.html) | 5 | Medium |
| EE-Norethindrone | Jevantique Lo | Oral | Tablet | [0.5MG-2.5](https://www.drugs.com/pro/jevantique-lo.html#s-34068-7) | 2 | Low |
| EE-Norethindrone | Jinteli | Oral | Tablet | [1MG-5MCG](https://dailymed.nlm.nih.gov/dailymed/drugInfo.cfm?setid=d667ca75-1a17-43fc-9077-f25ce0908f8d) | 5 | Medium |
| EE-Norethindrone | Norethindron-Ethinyl Estradiol | Oral | Tablet | [0.5MG-2.5](https://dailymed.nlm.nih.gov/dailymed/drugInfo.cfm?setid=cf549b5a-2707-4020-9f94-dab1260e1664) | 2 | Low |
| EE-Norethindrone | Norethindron-Ethinyl Estradiol | Oral | Tablet | [1MG-5MCG](https://dailymed.nlm.nih.gov/dailymed/drugInfo.cfm?setid=cf549b5a-2707-4020-9f94-dab1260e1664) | 5 | Medium |
| Medroxyprogesterone Acetate | Medroxyprogesterone Acetate | Oral | Tablet | [10MG](https://dailymed.nlm.nih.gov/dailymed/drugInfo.cfm?setid=2627eb11-06bf-4a45-9172-094468e3ca07) |  | NE |
| Medroxyprogesterone Acetate | Medroxyprogesterone Acetate | Oral | Tablet | [2.5MG](https://dailymed.nlm.nih.gov/dailymed/drugInfo.cfm?setid=2627eb11-06bf-4a45-9172-094468e3ca07) |  | NE |
| Medroxyprogesterone Acetate | Medroxyprogesterone Acetate | Oral | Tablet | [5MG](https://dailymed.nlm.nih.gov/dailymed/drugInfo.cfm?setid=2627eb11-06bf-4a45-9172-094468e3ca07) |  | NE |
| Medroxyprogesterone Acetate | Provera | Oral | Tablet | [10MG](https://dailymed.nlm.nih.gov/dailymed/drugInfo.cfm?setid=a586be28-96af-4fed-a13f-9b94fd4c7405) |  | NE |
| Medroxyprogesterone Acetate | Provera | Oral | Tablet | [2.5MG](https://dailymed.nlm.nih.gov/dailymed/drugInfo.cfm?setid=a586be28-96af-4fed-a13f-9b94fd4c7405) |  | NE |
| Medroxyprogesterone Acetate | Provera | Oral | Tablet | [5MG](https://dailymed.nlm.nih.gov/dailymed/drugInfo.cfm?setid=a586be28-96af-4fed-a13f-9b94fd4c7405) |  | NE |
| Progesterone | Progesterone | Oral | Capsule | [100MG](https://dailymed.nlm.nih.gov/dailymed/drugInfo.cfm?setid=d9e6ec7d-85ea-41a7-88ec-88cac22d5b7d) |  | NE |
| Progesterone | Progesterone | Oral | Capsule | [200MG](https://dailymed.nlm.nih.gov/dailymed/drugInfo.cfm?setid=d9e6ec7d-85ea-41a7-88ec-88cac22d5b7d) |  | NE |
| Progesterone | Prometrium | Oral | Capsule | [100MG](https://dailymed.nlm.nih.gov/dailymed/drugInfo.cfm?setid=a451784e-f6a0-414a-8409-3407f1d1d086) |  | NE |
| Progesterone | Prometrium | Oral | Capsule | [200MG](https://dailymed.nlm.nih.gov/dailymed/drugInfo.cfm?setid=a451784e-f6a0-414a-8409-3407f1d1d086) |  | NE |

**Notes:**

NE ^†^ = No estrogen ingredient. We only computed average daily doses for estrogen and for our primary analysis ignored differences in progestin ingredients, strengths and routes.

**S2 Table. List of CPT, ICD Diagnosis and Procedure codes used to define the absence of a uterus or hysterectomy surgery.**

|  | **ICD/CPT Codes** | **Description** | **# Claims** | **# Patients** |
| --- | --- | --- | --- | --- |
|  | All | any record of hysterectomy | 3,289,576 | 1,407,625 |
| ICD-9-CM Diagnosis | V8801 | Acquired absence of both cervix and uterus | 705,607 | 416,288 |
|  | V8802 | Acquired absence of uterus with remaining cervical stump | 9,901 | 7,679 |
| ICD-10-CM Diagnosis | Z90710 | Acquired absence of both cervix and uterus | 1,972,953 | 969,991 |
|  | Z90711 | Acquired absence of uterus with remaining cervical stump | 49,422 | 34,135 |
| ICD-9-CM Procedure | 6831 | Laparoscopic supracervical hysterectomy [LSH] | 2,039 | 2,036 |
|  | 6839 | Other and unspecified subtotal abdominal hysterectomy | 2,644 | 2,641 |
|  | 6841 | Laparoscopic total abdominal hysterectomy | 9,067 | 9,063 |
|  | 6849 | Other and unspecified total abdominal hysterectomy | 29,978 | 29,955 |
|  | 6851 | Laparoscopically assisted vaginal hysterectomy (LAVH) | 7,108 | 7,104 |
|  | 6859 | Other and unspecified vaginal hysterectomy | 17,320 | 17,303 |
|  | 6861 | Laparoscopic radical abdominal hysterectomy | 1,251 | 1,249 |
|  | 6869 | Other and unspecified radical abdominal hysterectomy | 1,845 | 1,844 |
|  | 6871 | Laparoscopic radical vaginal hysterectomy [LRVH] | 240 | 239 |
|  | 6879 | Other and unspecified radical vaginal hysterectomy | 121 | 121 |
| ICD-10-CM PCS | 0UT90ZL | Resection of Uterus, Supracervical, Open Approach | 1,073 | 1,070 |
|  | 0UT90ZZ | Resection of Uterus, Open Approach | 25,414 | 25,387 |
|  | 0UT94ZL | Resection of Uterus, Supracervical, Percutaneous Endoscopic Approach | 591 | 590 |
|  | 0UT94ZZ | Resection of Uterus, Percutaneous Endoscopic Approach | 8,132 | 8,121 |
|  | 0UT97ZL | Resection of Uterus, Supracervical, Via Natural or Artificial Opening | 10 | 10 |
|  | 0UT97ZZ | Resection of Uterus, Via Natural or Artificial Opening | 4,919 | 4,914 |
|  | 0UT98ZL | Resection of Uterus, Supracervical, Via Natural or Artificial Opening Endoscopic | 17 | 17 |
|  | 0UT98ZZ | Resection of Uterus, Via Natural or Artificial Opening Endoscopic | 341 | 341 |
|  | 0UT9FZL | Resection of Uterus, Supracervical, Via Natural or Artificial Opening With Percutaneous Endoscopic Assistance | 53 | 53 |
|  | 0UT9FZZ | Resection of Uterus, Via Natural or Artificial Opening With Percutaneous Endoscopic Assistance | 8,132 | 8,121 |
|  | 0UTC0ZZ | Resection of Cervix, Open Approach | 13,493 | 13,487 |
|  | 0UTC4ZZ | Resection of Cervix, Percutaneous Endoscopic Approach | 4,199 | 4,192 |
|  | 0UTC7ZZ | Resection of Cervix, Via Natural or Artificial Opening | 3,692 | 3,692 |
|  | 0UTC8ZZ | Resection of Cervix, Via Natural or Artificial Opening Endoscopic | 409 | 408 |
| CPT - Abdominal hysterectomy; includes that by laparoscope | 58150 | Total abdominal hysterectomy (corpus and cervix), with or without removal of tube(s), with or without removal of ovary(s) | 37,548 | 26,909 |
|  | 58152 | Total abdominal hysterectomy (corpus and cervix), with or without removal of tube(s), with or without removal of ovary(s); with colpo-urethrocystopexy (eg, Marshall-Marchetti-Krantz, Burch) | 860 | 583 |
|  | 58180 | Supracervical abdominal hysterectomy (subtotal hysterectomy), with or without removal of tube(s), with or without removal of ovary(s) | 4,591 | 3,357 |
|  | 58200 | Total abdominal hysterectomy, including partial vaginectomy, with para-aortic and pelvic lymph node sampling, with or without removal of tube(s), with or without removal of ovary(s) | 2,993 | 2,395 |
|  | 58210 | Radical abdominal hysterectomy, with bilateral total pelvic lymphadenectomy and para-aortic lymph node sampling (biopsy), with or without removal of tube(s), with or without removal of ovary(s) | 6,984 | 5,394 |
|  | 58240 | Pelvic exenteration for gynecologic malignancy, with total abdominal hysterectomy or cervicectomy, with or without removal of tube(s), with or without removal of ovary(s), with removal of bladder and ureteral transplantations, and/or abdominoperineal resection of rectum and colon and colostomy, or any combination thereof | 1,046 | 666 |
|  | 58541 | Laparoscopy, surgical, supracervical hysterectomy, for uterus 250 g or less | 2,754 | 1,726 |
|  | 58542 | Laparoscopy, surgical, supracervical hysterectomy, for uterus 250 g or less; with removal of tube(s) and/or ovary(s) | 23,276 | 12,111 |
|  | 58543 | Laparoscopy, surgical, supracervical hysterectomy, for uterus greater than 250 g | 293 | 219 |
|  | 58544 | Laparoscopy, surgical, supracervical hysterectomy, for uterus greater than 250 g; with removal of tube(s) and/or ovary(s) | 825 | 561 |
|  | 58548 | Laparoscopy, surgical, with radical hysterectomy, with bilateral total pelvic lymphadenectomy and para-aortic lymph node sampling (biopsy), with removal of tube(s) and ovary(s), if performed | 12,113 | 8,945 |
|  | 58550 | Laparoscopy, surgical, with vaginal hysterectomy, for uterus 250 g or less | 3,613 | 2,482 |
|  | 58552 | Laparoscopy, surgical, with vaginal hysterectomy, for uterus 250 g or less; with removal of tubes(s) and /or ovary(s) | 45,405 | 26,926 |
|  | 58553 | Laparoscopy, surgical, with vaginal hysterectomy, for uterus greater than 250 g | 251 | 192 |
|  | 58554 | Laparoscopy, surgical, with vaginal hysterectomy, for uterus greater than 250 g; with removal of tube(s) and/or ovary(s) | 2,047 | 1,472 |
|  | 58570 | Laparoscopy, surgical, with total hysterectomy, for uterus 250 g or less | 3,773 | 2,603 |
|  | 58571 | Laparoscopy, surgical, with total hysterectomy, for uterus 250 g or less; with removal of tube(s) and/or ovary(s) | 147,913 | 77,995 |
|  | 58572 | Laparoscopy, surgical, with total hysterectomy, for uterus greater than 250 g | 671 | 518 |
|  | 58573 | Laparoscopy, surgical, with total hysterectomy, for uterus greater than 250 g; with removal of tube(s) and/or ovary(s) | 9,761 | 6,102 |
|  | 58575 |  | 1,538 | 1,133 |
|  | 58951 | Resection (initial) of ovarian, tubal or primary peritoneal malignancy with bilateral salpingo-oophorectomy and omentectomy; with total abdominal hysterectomy, pelvic and limited para-aortic lymphadenectomy | 3,156 | 2,512 |
|  | 58953 | Bilateral salpingo-oophorectomy with omentectomy, total abdominal hysterectomy and radical dissection for debulking | 8,405 | 6,593 |
|  | 58954 | Bilateral salpingo-oophorectomy with omentectomy, total abdominal hysterectomy and radical dissection for debulking; with pelvic lymphadenectomy and limited para-aortic lymphadenectomy | 6,499 | 4,937 |
|  | 58956 | Bilateral salpingo-oophorectomy with total omentectomy, total abdominal hysterectomy for malignancy | 2,512 | 1,988 |
|  | 59525 | Subtotal or total hysterectomy after cesarean delivery | - | - |
|  | 58575 | Laparoscopy, surgical, total hysterectomy for resection of malignancy (tumor debulking), with omentectomy including salpingo-oophorectomy, unilateral or bilateral, when performed | - | - |
| CPT - Vaginal hysterectomy | 51925 | Closure of vesicouterine fistula; with hysterectomy | 4 | 3 |
|  | 58260 | Vaginal hysterectomy, for uterus 250 g or less | 55,354 | 29,800 |
|  | 58262 | Vaginal hysterectomy, for uterus 250 g or less; with removal of tube(s), and/or ovary(s) | 37,492 | 20,639 |
|  | 58263 | Vaginal hysterectomy, for uterus 250 g or less; with removal of tube(s), and/or ovary(s), with repair of enterocele | 6,333 | 4,486 |
|  | 58267 | Vaginal hysterectomy, for uterus 250 g or less; with colpo-urethrocystopexy (Marshall-Marchetti-Krantz type, Pereyra type) with or without endoscopic control | 767 | 531 |
|  | 58270 | Vaginal hysterectomy, for uterus 250 g or less; with repair of enterocele | 5,785 | 4,011 |
|  | 58275 | Vaginal hysterectomy, for uterus 250 g or less; | 577 | 455 |
|  | 58280 | Vaginal hysterectomy, with total or partial vaginectomy; with repair of enterocele | 338 | 262 |
|  | 58285 | Vaginal hysterectomy, radical (Schauta type operation) | 70 | 56 |
|  | 58290 | Vaginal hysterectomy, for uterus greater than 250 g | 602 | 414 |
|  | 58291 | Vaginal hysterectomy, for uterus greater than 250 g; with removal of tube(s) and/or ovary(s) | 644 | 449 |
|  | 58292 | Vaginal hysterectomy, for uterus greater than 250 g; with removal of tube(s) and/or ovary(s), with repair of enterocele | 157 | 118 |
|  | 58293 | Vaginal hysterectomy, for uterus greater than 250 g; with colpo-urethrocystopexy (Marshall-Marchetti-Krantz type, Pereyra type) with or without endoscopic control | 53 | 39 |
|  | 58294 | Vaginal hysterectomy, for uterus greater than 250 g; with repair of enterocele | 127 | 95 |
